# High prevalence of SARS-CoV-2 swab positivity in England during September 2020: interim report of round 5 of REACT-1 study

**DOI:** 10.1101/2020.09.30.20204727

**Authors:** Steven Riley, Kylie E. C. Ainslie, Oliver Eales, Caroline E. Walters, Haowei Wang, Christina Atchison, Claudio Fronterre, Peter J. Diggle, Deborah Ashby, Christl A. Donnelly, Graham Cooke, Wendy Barclay, Helen Ward, Ara Darzi, Paul Elliott

## Abstract

**Background:** REACT-1 is a community survey of PCR confirmed swab-positivity for SARS-CoV-2 among random samples of the population in England. This interim report includes data from the fifth round of data collection currently underway for swabs sampled from the 18th to 26th September 2020.

**Methods:** Repeated cross-sectional surveys of random samples of the population aged 5 years and over in England with sample size ranging from 120,000 to 160,000 people in each round of data collection. Collection of self-administered nose and throat swab for PCR and questionnaire data. Prevalence of swab-positivity by round and by demographic variables including age, sex, region, ethnicity. Estimation of reproduction number (R) between and within rounds, and time trends using exponential growth or decay model. Assessment of geographical clustering based on boundary-free spatial model.

**Results:** Over the 9 days for which data are available, we find 363 positives from 84,610 samples giving a weighted prevalence to date of 0.55% (0.47%, 0.64%) in round 5. This implies that 411,000 (351,000, 478,000) people in England are virus-positive under the assumption that the swab assay is 75% sensitive. Using data from the most recent two rounds, we estimate a doubling time of 10.6 (9.4, 12.0) days covering the period 20th August to 26th September, corresponding to a reproduction number R of 1.47 (1.40, 1.53). Using data only from round 5 we estimate a reproduction number of 1.06 (0.74, 1.46) with probability of 63% that R is greater than 1. Between rounds 4 and 5 there was a marked increase in unweighted prevalence at all ages. In the most recent data, prevalence was highest in the 18 to 24 yrs age group at 0.96% (0.68%, 1.36%). At 65+ yrs prevalence increased ∼7-fold between rounds 4 and 5 from 0.04% (0.03%, 0.07%) to 0.29% (0.23%, 0.37%). Prevalence increased in all regions between rounds 4 and 5, giving the highest unweighted prevalence in round 5 in the North West at 0.86% (0.69%, 1.06%). In London, prevalence increased ∼5-fold from 0.10% (0.06%, 0.17%) to 0.49% (0.36%, 0.68%). Regional R values ranged from 1.32 (1.16,1.50) in Yorkshire and the Humber to 1.63 (1.42, 1.88) in the East Midlands over the same period. In the most recent data, there was extensive clustering in the North West, Midlands and in and around London with pockets of clustering in other regions including the South West, North East and East of England. Odds of swab-positivity were ∼2-fold higher in people of Asian and Black ethnicity compared with white participants.

**Conclusion:** Rapid growth has led to high prevalence of SARS-CoV-2 virus in England among all regions and age groups, including those age groups at highest risk. Although there is evidence of a recent deceleration in the epidemic, current levels of prevalence will inevitably result in additional hospitalisations and mortality in coming weeks. A re-doubling of public health efforts is needed to return to a declining phase of the epidemic.

## Introduction

England is experiencing one of the largest epidemics of COVID-19 in Europe with substantial numbers of hospitalisations and mortality during the first wave of infections that peaked in March and April 2020 [1]. A national lockdown was implemented on 23 March 2020 and continued through May 2020. We established the REal-time Assessment of Community Transmission-1 (REACT-1) study to track prevalence of SARS-CoV-2 virus in the community across England, starting in May 2020 as England exited lockdown and continuing with repeated representative surveys to present [2–4]. After large falls in prevalence from May through to early August 2020 we detected an upturn in mid August to early September 2020, with reproduction number (R) value estimated to be 1.7 (1.4, 2.0). We report here interim results from the most recent round of data collection from 18th to 26th September 2020 and compare prevalence and trends with the previous four rounds.

## Methods

REACT-1 is quantifying the prevalence of SARS-CoV-2 in the community in England [2] based on obtaining repeated random samples of the population at ages five years and over using the National Health Service (NHS) list of patients registered with a general practitioner. Sampling was stratified by the 315 lower-tier local authorities in England. People were invited to take part by post and asked to complete a registration questionnaire. A self-administered swab kit was then posted to the individual with instructions for completing the swab, including online video (parent/guardian administered from ages 5 to 12 years). Participants also completed an online or telephone questionnaire. Swabs were refrigerated at home then picked up from the participant’s home and sent on a cold chain to the laboratory for testing using PCR with two gene targets (E gene and N gene). Cycle threshold (CT) values were used as a proxy for intensity of viral load. A PCR test was considered positive if both gene targets were detected or if N gene was detected with CT value less than 37 [3].

Data collection has so far been completed over four rounds starting 1 May 2020, involving between 120,000 and 160,000 people at each round, with a further (fifth) round of data collection currently underway (Table 1). This report summarises data from the first four rounds and provides interim data for round 5.

**Table 1.** Weighted and unweighted prevalence of swab-positivity across four completed rounds of the REACT-1 study and the partially complete round 5.

| Round | Tested swabs | Positive swabs | Unweighted prevalence (95% CI) | Weighted prevalence (95% CI) | First sample | Last sample |
| --- | --- | --- | --- | --- | --- | --- |
| 1 | 120,620 | 159 | 0.13% (0.11%, 0.15%) | 0.16% (0.13%, 0.19%) | 1/5/2020 | 1/6/2020 |
| 2 | 159,199 | 123 | 0.077% (0.065%, 0.092%) | 0.088% (0.068%, 0.11%) | 19/6/2020 | 7/7/2020 |
| 3 | 161,560 | 54 | 0.033% (0.025%, 0.043%) | 0.040% (0.027%, 0.053%) | 24/7/2020 | 11/8/2020 |
| 4 | 154,325 | 137 | 0.089% (0.075%, 0.11%) | 0.13% (0.096%, 0.15%) | 20/8/2020 | 8/9/2020 |
| 5* | 84,610 | 363 | 0.43% (0.39%, 0.48%) | 0.55% (0.47%, 0.64%) | 18/9/2020 | 26/9/2020 |

\* Not yet complete

### Analyses

We estimated prevalence by dividing numbers of swab positive results by numbers tested. We corrected for the sampling design and differential response by reweighting the overall prevalence estimate according to the population of England taking into account age, sex, region, ethnicity and deprivation; all other prevalence estimates were unweighted. We used an exponential model to investigate trends over time, with the assumption that the number of positive swabs each day arose from a binomial distribution, based on day of swabbing or, if unavailable, day of collection. We used a bivariate no-u-turn sampler with uniform prior distributions to calculate posterior credible intervals [5]. We estimated the reproduction number R by assuming the generation time followed a gamma distribution (mean 6.29 days, standard deviation of 4.2) [6]. R was estimated across sequential rounds and separately for each round. We carried out logistic regression (adjusted for age, sex, region, key worker status, ethnicity, and household size) to obtain odds ratio estimates and 95% confidence intervals for associations of swab positivity with covariates.

We investigated possible clustering of cases across rounds 1 and 2, rounds 2 and 3, rounds 3 and 4, and interim data for round 5. We first calculated distances (up to 30 km) between home locations of swab-positive participants with available georeferencing information; as a control, we randomly sampled 5,000 times the same number of swab-negative as swab-positive participants and calculated the equivalent distances. We then located individuals who appeared frequently within nearby pairs. From the negative pairs, we obtained a null cumulative distribution of frequencies by which participants appeared in pairs within 30 km of each other and obtained the corresponding single cumulative frequency curve for swab-positive participants. A cluster was declared if a participant appeared in nearby pairs more times than the point at which the swab-positive cumulative curve diverged from the central 90% region of the swab-negative distribution. We then calculated the area of the convex hull which contained the home locations for the clustered swab-positive participants and 10,000 samples of an identical number of swab-negative participants.

Research ethics approval was obtained from the South Central-Berkshire B Research Ethics Committee (IRAS ID: 283787).

## Results

To date over 680,000 swabs have been tested in the REACT-1 study (Table 1). Over the 9 days for which data are available in the most recent round, we find 363 positives from 84,610 samples for a weighted prevalence of 0.55% (0.47%, 0.64%), which continues the upwards trend in prevalence seen in the previous round (Table 1, Figure 1). This is the highest observed prevalence since the beginning of the study in May 2020 and more than a four-fold increase in the weighted prevalence observed during round 4 (Table 1). Using data from rounds 4 and 5 and we estimate a doubling time of 10.6 (9.4, 12.0) days covering the period 20th August to 26th September 2020 using a model of exponential growth and decay. This corresponds to a reproduction number R of 1.47 (1.40, 1.53) (Table 2, Figure 2A). However, using only the most recent data we observe a more gradual slope (Figure 2B), with an estimated reproduction number of 1.06 (0.74, 1.46) and 63% probability that R is greater than 1 (Table 2). We tested the sensitivity of our estimates of R to subsets of the data. Our results were consistent when using: positives who did not report symptoms, positives on both gene targets, and positives on both genes or with CT values for N gene less than 35.

**Table 2.** Estimates of growth rate and reproduction number for rounds 4 (complete) and 5 (partial).

| Round | Round | Growth rate (r) | Reproduction number | P(R>1) |
| --- | --- | --- | --- | --- |
| All positives | 4 and 5 | 0.065 ( 0.058 , 0.074 ) | 1.47 ( 1.4 , 1.53 ) | 1.00 |
|  | 5 | 0.01 ( -0.045 , 0.064 ) | 1.06 ( 0.74 , 1.46 ) | 0.63 |
| Asymptomatic positives | 4 and 5 | 0.055 ( 0.043 , 0.066 ) | 1.38 ( 1.3 , 1.47 ) | 1.00 |
|  | 5 | -0.012 ( -0.095 , 0.071 ) | 0.92 ( 0.5 , 1.51 ) | 0.38 |
| Positive for both gene targets | 4 and 5 | 0.074 ( 0.065 , 0.085 ) | 1.54 ( 1.46 , 1.62 ) | 1.00 |
|  | 5 | -0.025 ( -0.089 , 0.039 ) | 0.85 ( 0.52 , 1.27 ) | 0.21 |
| Positive for both gene targets or N gene CT < 35* | 4 and 5 | 0.068 ( 0.059 , 0.077 ) | 1.49 ( 1.42 , 1.56 ) | 1.00 |
|  | 5 | -0.006 ( -0.065 , 0.052 ) | 0.97 ( 0.64 , 1.36 ) | 0.42 |

\* All positives use a CT threshold of 37

**Figure 1.**
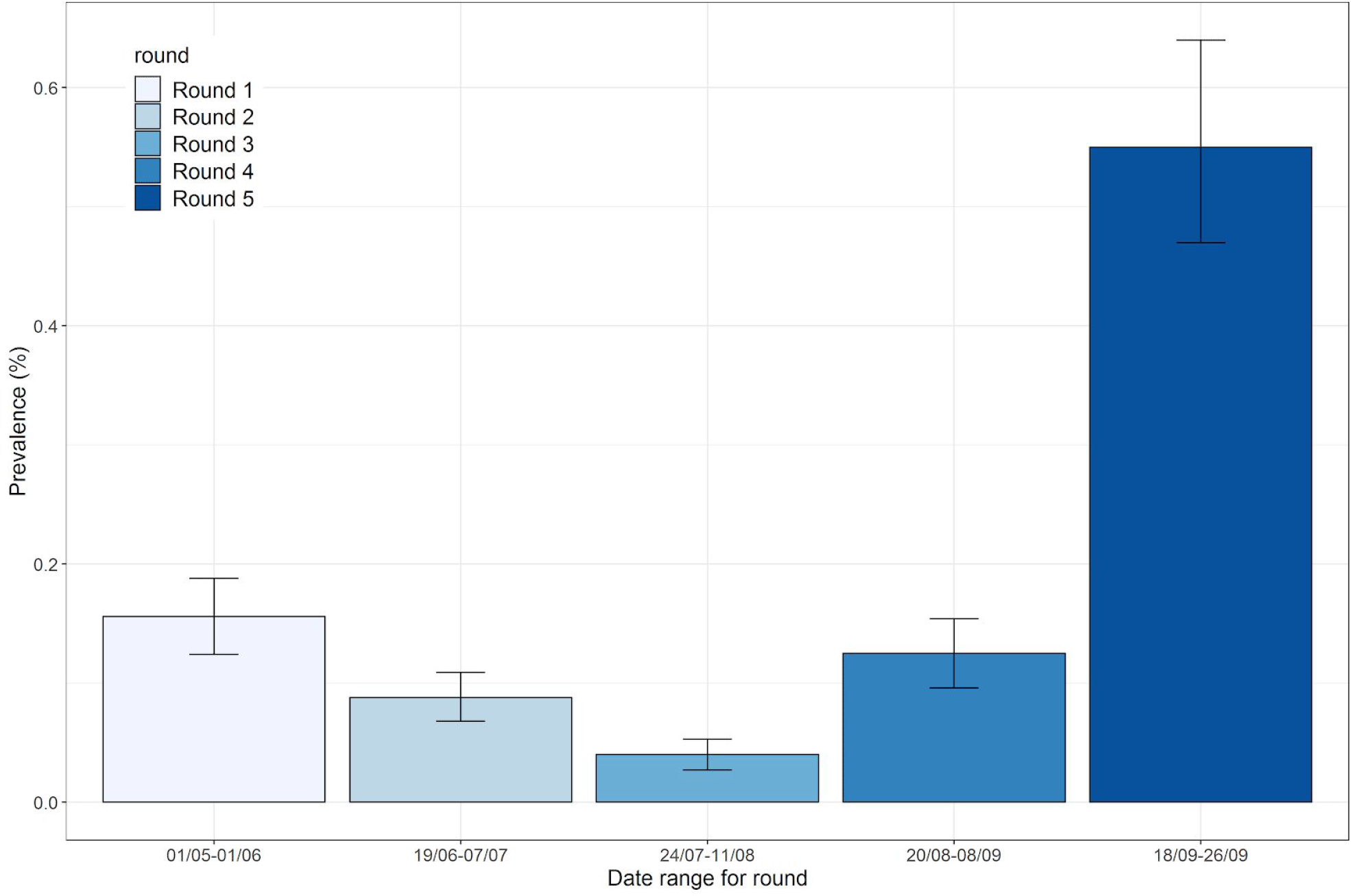
Weighted prevalence of swab-positivity in England for completed rounds 1 to 4 and partially complete round 5 of the REACT-1 study. Error bars show 95% binomial confidence bounds.

**Figure 2.**
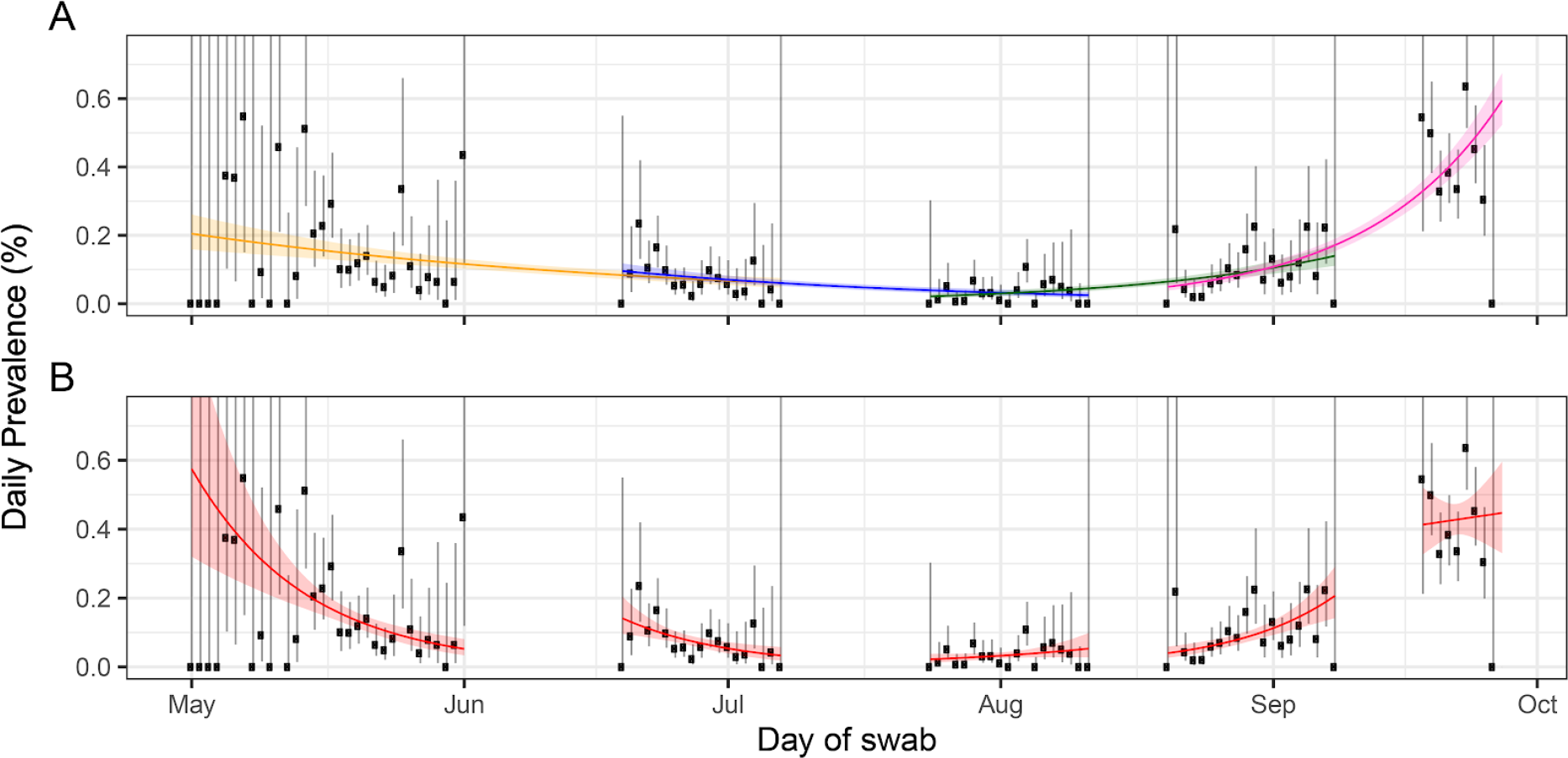
Constant growth rate models fit to REACT-1 data for sequential and individual rounds. **A** models fit to REACT-1 data for sequential rounds; 1 and 2 (yellow), 2 and 3 (blue), 3 and 4 (green) and 4 and 5 (pink). **B** models fit to individual rounds only (red). Vertical lines show 95% confidence intervals for observed prevalence (black points). Shaded regions show 95% posterior credibility intervals for growth models.

Between rounds 4 and 5 there was a marked increase in unweighted prevalence at all ages (Figure 3, Table S2). In the most recent data, prevalence was highest in the 18 to 24 yrs age group at 0.96% (0.68%, 1.36%), while at ages 65+ yrs prevalence increased ∼7-fold between rounds 4 and 5 from 0.04% (0.03%, 0.07%) to 0.29% (0.23%, 0.37%). Overall, the age pattern of prevalence was consistent with round 4. Odds of infection were ∼2-fold higher in people of Asian and Black ethnicity compared with white participants (Table S2, Figure S2).

**Figure 3.**
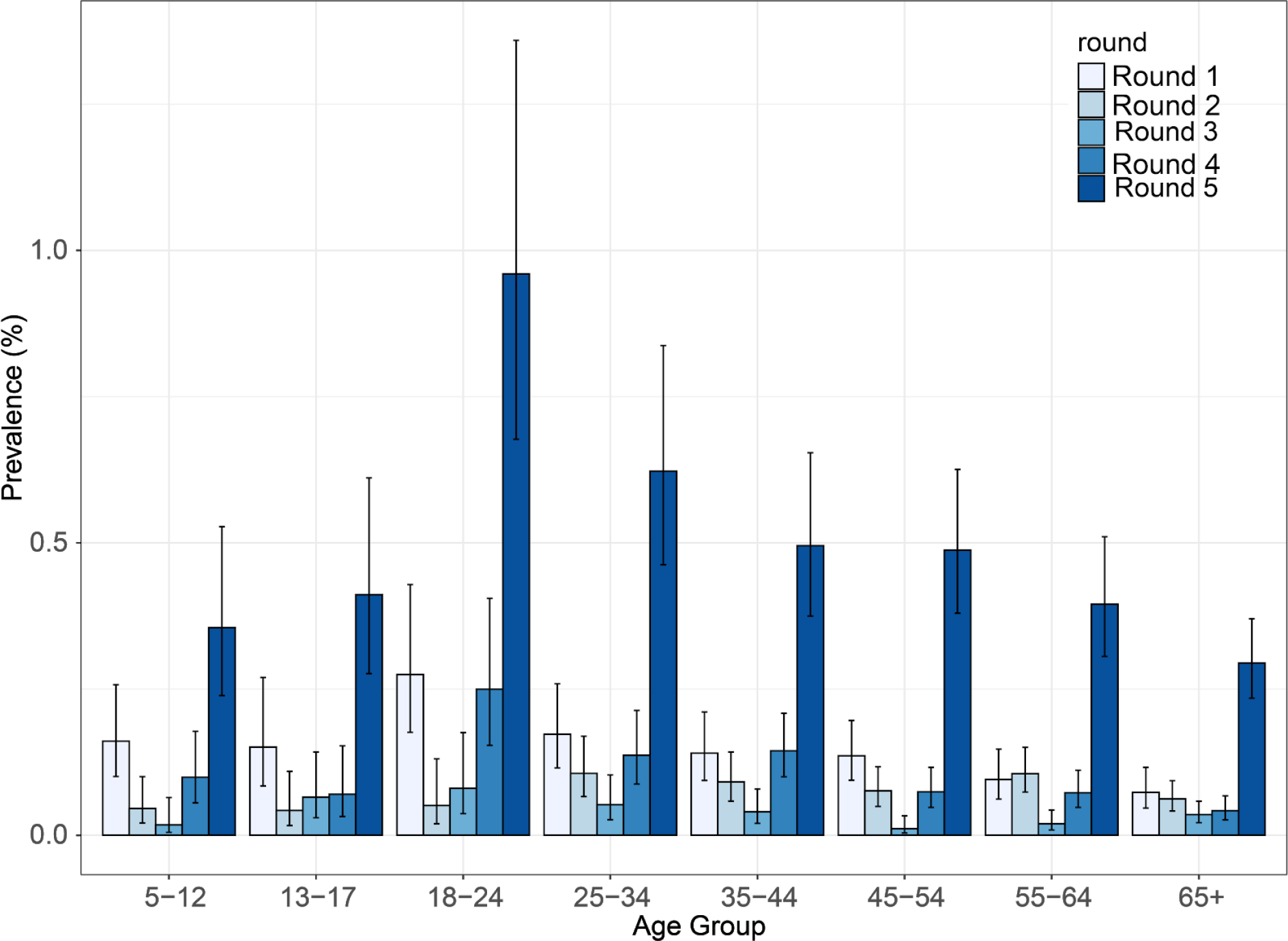
Unweighted prevalence of swab positivity by age by round.

Prevalence increased in all regions between rounds 4 and 5 (Figures 4 and 5, Table S2) with highest unweighted prevalence in round 5 in the North West at 0.86% (0.69%, 1.06%). In London, prevalence increased ∼5-fold from 0.10% (0.06%, 0.17%) to 0.49% (0.36%, 0.68%), with R estimates ranging from 1.32 (1.16,1.50) in Yorkshire and The Humber to 1.63 (1.42, 1.88) in the East Midlands over the same period (Figure S1, Table S1). Extensive clustering was evident in the North West, Midlands and in and around London (Figure 6), with new pockets of clustering occurring in other regions including the South West, North East and East of England.

**Figure 4.**
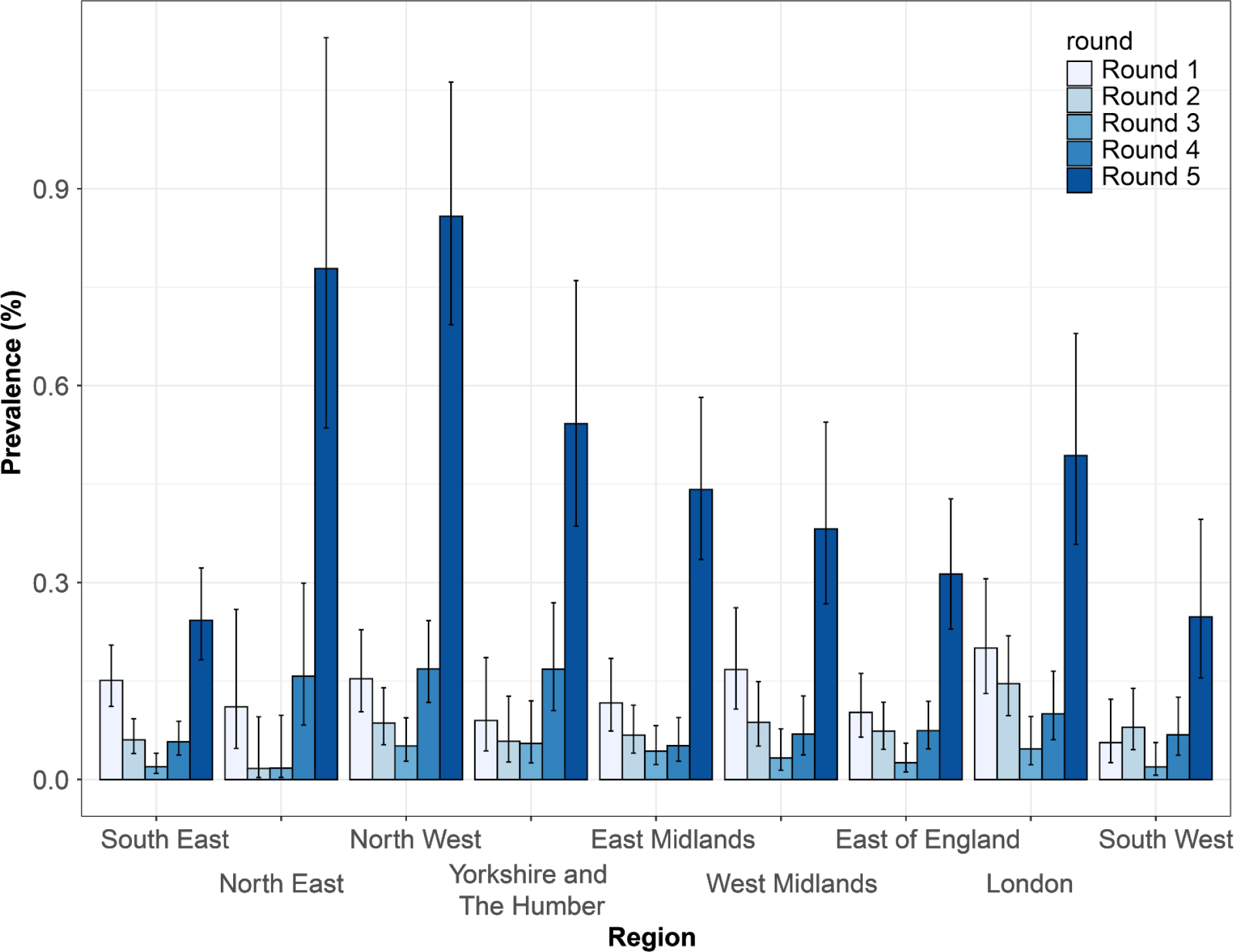
Unweighted prevalence of swab positivity by region by round.

**Figure 5.**
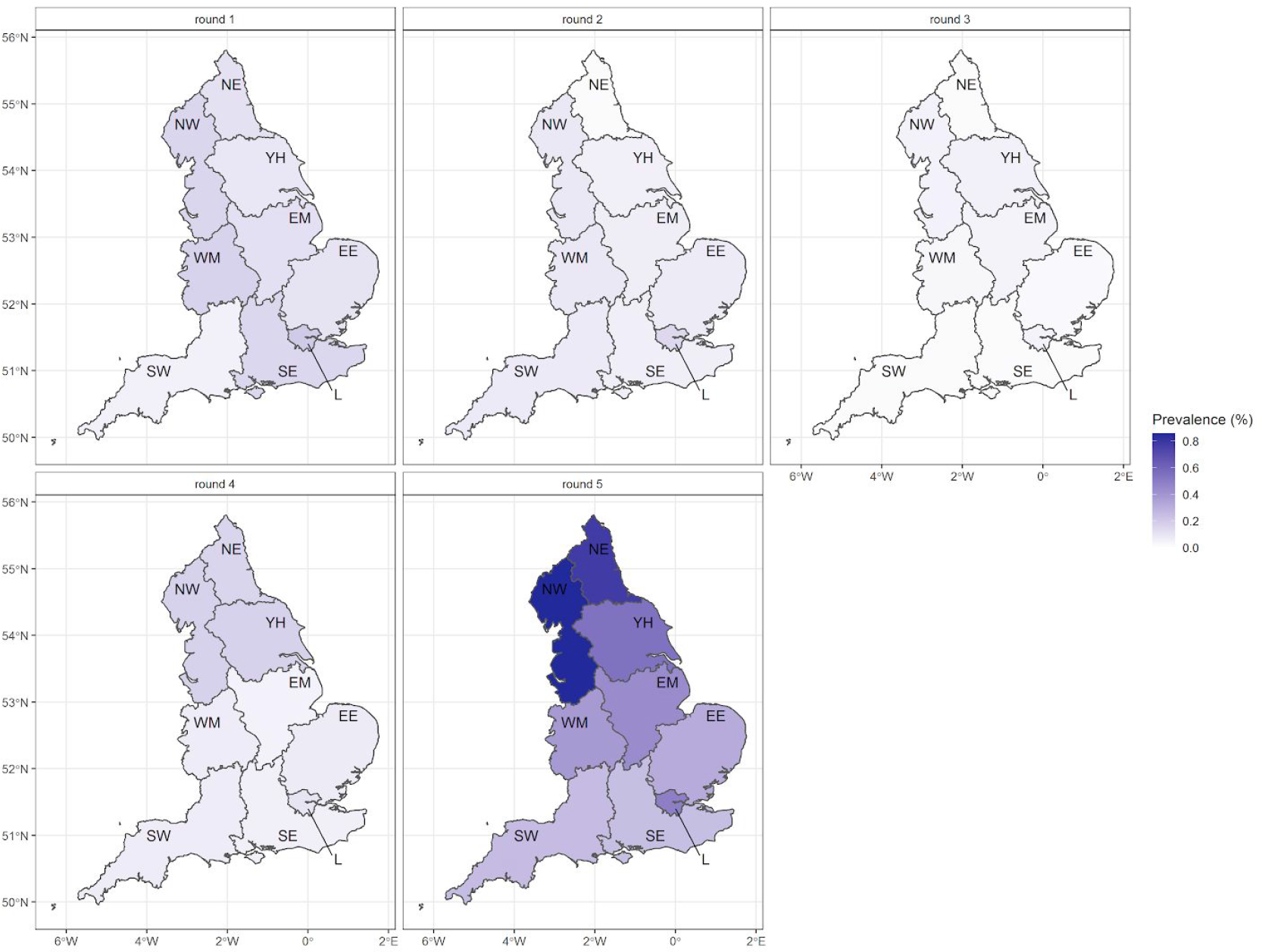
Unweighted prevalence by England region by round. NE = North East, NW = North West, YH = Yorkshire and The Humber, EM = East Midlands, WM = West Midlands, EE = East of England, L = London, SE = South East, SW = South West.

**Figure 6.**
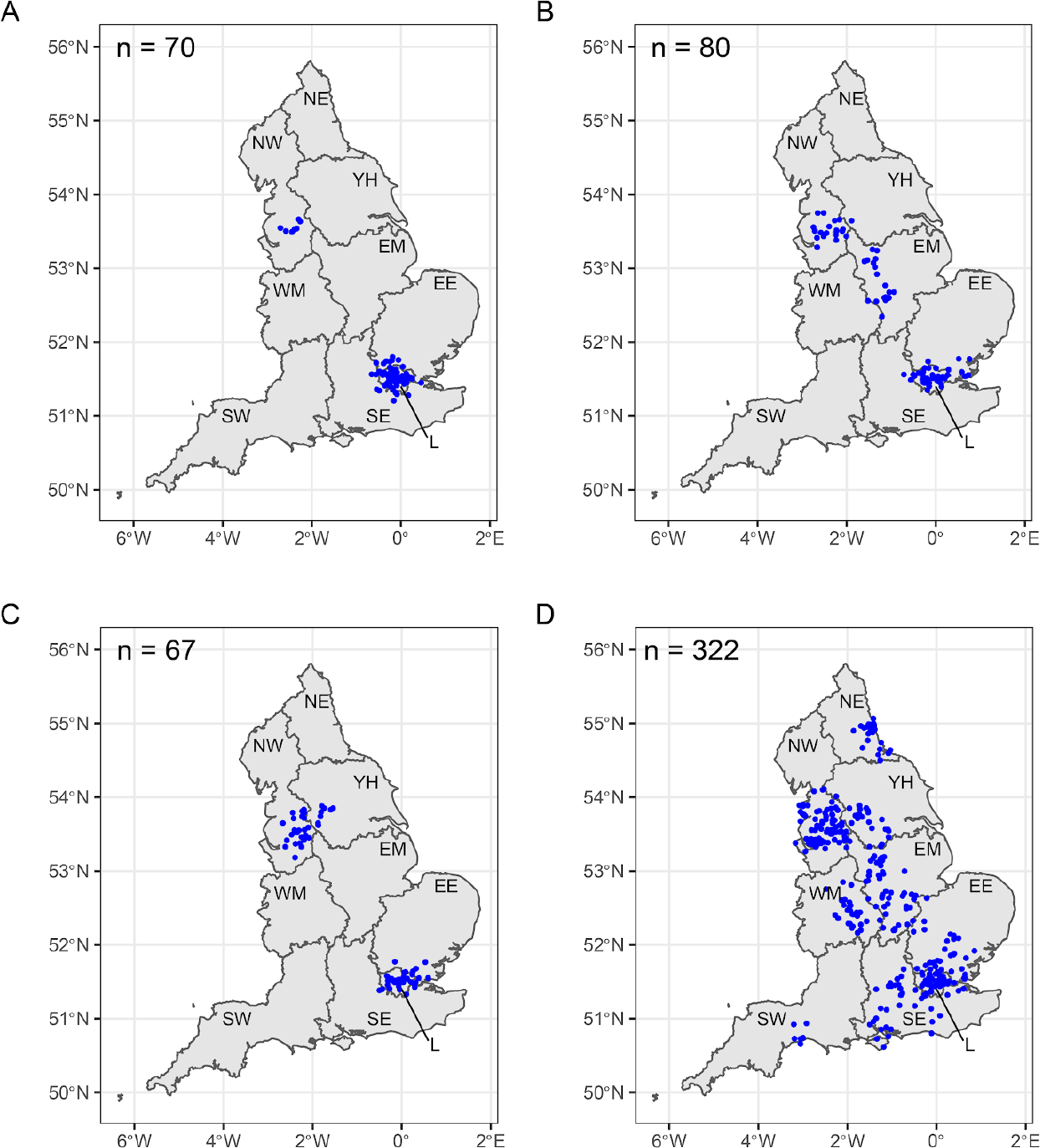
A (rounds 1 and 2) and **B** (rounds 2 and 3) **C** (rounds 3 and 4) and **D** (round 5) jittered home locations of swab-positive participants who most frequently formed close pairs with other swab-positive participants. Regions of England indicated by: L, London; SE, South East; SW, South West; WM, West Midlands; EE, East of England; EM, East Midlands; NW, North West; YH, Yorkshire and The Humber; and NE, North East.

## Discussion

In this interim report from round 5 of the REACT-1 study to 26 September 2020, we found prevalence of swab positivity had increased to over 1 in 200 across the population in England. It was highest in the 18-24 year old group with seven-fold increase at ages 65 and above from late August/early September 2020. All regions of the country showed a rise with highest prevalence in the North West and five-fold increase in prevalence in London over this period. We also found evidence of widely distributed local clustering of swab positive individuals. However, there was some indication in recent data that the rate of increase in prevalence may have slowed.

The REACT-1 programme has been tracking the spread of SARS-CoV-2 in the community in England since May 2020 when national lockdown was still in place. We showed rapid declines in prevalence of swab positivity based on PCR during May to mid-August 2020, when prevalence was at its lowest. We then detected a rise in prevalence from end of August to early September 2020, with R reliably above one and a doubling time of seven to eight days [4]. Since then prevalence has increased such that we estimate there to be 411,000 (351,000, 478,000) people in England who are virus-positive on a given day assuming a sensitivity of 75% for a nose and throat swab conducted at home [7].

The data on prevalence by age indicate that the recent resurgence in infections is not limited to the 18-24 year old group, where prevalence of infections is now around one percent, but extends across the whole age range. Importantly, those at older ages who are at most risk from SARS-CoV-2 infection have seen the largest relative rise in infections in recent data. These infections will have an unavoidable impact on hospitalisations and mortality, since a proportion of such cases are likely to result in severe infections [8]. The high rates of infection in young adults have implications for the spread of the virus in colleges and universities where local outbreaks are already being detected [9].

The rise in prevalence that we detected in September is broadly consistent with other data that also found a rise in the numbers of cases [10] including individuals testing positive through the government’s ‘test and trace’ programme [11]. However, unlike symptomatic testing programmes like ‘test and trace’, our study is not dependent on the extent to which symptomatic individuals present for testing nor on issues of service capacity. As our study is based on random samples of the general population, the REACT-1 programme is able to provide reliable and timely data on trends in prevalence of SARS-CoV-2 that are less affected by such biases.

The slowing of the rate of increase in prevalence seen in the most recent data is also consistent with other data sources including routine testing through ‘test and trace’ and calls to the National Health Service and emergency services [11]. This follows a reinforced public health messaging campaign by the government for individuals to comply with social distancing measures, restrict numbers of people visiting private households or meeting outdoors at any one time, and, more recently, closure of pubs and bars by 10 pm and enforcement of social distancing rules [12]. A series of local lockdowns has also been implemented that may have contained spread of the virus in communities thought to be at highest risk of transmission. It is plausible that these measures are increasing both public awareness of the current scale of the epidemic in England and compliance with social distancing rules and other measures, such as hand-washing and wearing of face covers; these in turn may have fed through to a lowering of the R value. Notwithstanding these recent trends, the current level of infections, particularly at older ages, will result in additional hospital admissions and mortality from COVID-19 in the coming weeks.

The regional data suggest that the recent increases in prevalence of SARS-CoV-2 have been widespread and not restricted to local outbreaks, although we also found evidence of local clustering. The most recent data indicate large increases in prevalence in the North of England, with evidence also for large increases in London and the Midlands, but less so in the South East and South West. This implies that reinforcement of public health measures on social mixing and distancing needs to occur at the national level, and not only at the local level where ‘hotspots’ are detected. Recent government initiatives have adopted this dual approach.

As we reported previously [13] the recent data no longer show that the epidemic in England is being driven by outbreaks in hospitals or care homes with consequent high rates in health care and care home workers [3], but now reflects predominantly community transmission. We also reported previously higher swab positivity rates in minority ethnic groups and this has continued, with rates twice as high in people of Asian and Black ethnicities compared with white people.

Limitations of our study include possible differential responses to taking part, although in our estimates of prevalence overall we were able to correct for key differences between the population characteristics of our sample and of England as a whole. Also, it is possible that some of the recent rise in prevalence may be explained by difficulties in obtaining a PCR test through the routine testing programme. This is supported by the prevalence of people without symptoms on the day of testing or week before being lower in the most recent round compared to previous rounds. However, this increase in the proportion of swab-positive participants with symptoms may also reflect higher viral loads in the population (which may be more likely to be associated with symptoms) and this is supported by a reduction in CT values in the most recent data (not shown). Furthermore, our rates of growth in prevalence of the virus were similar among symptomatic people and those without symptoms who would be largely ineligible for testing through the routine ‘test and trace’ programme. An additional limitation is that a nose and throat swab may have limited sensitivity (∼70% to 80%) [7] to detect virus, but this should not affect trends in prevalence within or between rounds.

Our study is being undertaken at a critical time in the progression of the SARS-CoV-2 epidemic in England as we enter a second wave. The initial lockdown in March 2020 and restrictions of population and individual behaviours in the subsequent three months resulted in marked reductions in prevalence of the virus to low levels by the beginning of August. However, since mid-August when we first detected a rise in prevalence, there has been a resurgence of the virus in the community, with rates higher now than at any time since we started measuring prevalence in May 2020. Hence there is an urgent need for a redoubling of public health efforts to reduce transmission of the virus in the community. The primary aim should be to limit hospitalisations and deaths from COVID-19 that will inevitably follow the current high prevalence of SARS-CoV-2 infection in the community.

## Data Availability

The original datasets generated or analysed, or both, during this study are not publicly available because of governance restrictions and the identifiable nature of the data.

## Acknowledgements

SR, CAD acknowledge support: MRC Centre for Global Infectious Disease Analysis, National Institute for Health Research (NIHR) Health Protection Research Unit (HPRU), Wellcome Trust (200861/Z/16/Z, 200187/Z/15/Z), and Centres for Disease Control and Prevention (US, U01CK0005-01-02). GC is supported by an NIHR Professorship. PE is Director of the MRC Centre for Environment and Health (MR/L01341X/1, MR/S019669/1). PE acknowledges support from the NIHR Imperial Biomedical Research Centre and the NIHR HPRUs in Environmental Exposures and Health and Chemical and Radiation Threats and Hazards, the British Heart Foundation Centre for Research Excellence at Imperial College London (RE/18/4/34215) and the UK Dementia Research Institute at Imperial (MC_PC_17114). We thank The Huo Family Foundation for their support of our work on COVID-19.

We thank key collaborators on this work -- Ipsos MORI: Kelly Beaver, Sam Clemens, Gary Welch, Nick Gilby, Andrew Cleary and Kelly Ward; Institute of Global Health Innovation at Imperial College: Gianluca Fontana, Dr Hutan Ashrafian, Sutha Satkunarajah and Lenny Naar; MRC Centre for Environment and Health, Imperial College London: Daniela Fecht; North West London Pathology and Public Health England for help in calibration of the laboratory analyses; NHS Digital for access to the NHS register; and the Department of Health and Social Care for logistic support. SR acknowledges helpful discussion with members of the UK Government Office for Science (GO-Science) Scientific Pandemic Influenza – Modelling (SPI-M) committee.

## Main tables and figures

Tables and data for Figure 2 available electronically [here](https://docs.google.com/spreadsheets/d/14B-cMBlgfQgmnPP0XK9iRB2nsxQltHLlcyyqVEGXKs4/edit?usp=sharing).

## Supporting tables and figures

**Table S1.** Estimates of growth rate and reproduction number for regions for rounds 4 (complete) and 5 (partial).

| Region | Growth rate (r) | Reproduction number | P(R>1) | Doubling time |
| --- | --- | --- | --- | --- |
| East Midlands | 0.086 ( 0.060 , 0.114 ) | 1.63 ( 1.42 , 1.88 ) | 1 | 8.1 ( 6.1 , 11.6 ) |
| West Midlands | 0.071 ( 0.043 , 0.103 ) | 1.51 ( 1.30 , 1.78 ) | 1 | 9.8 ( 6.7 , 16.0 ) |
| East of England | 0.060 ( 0.037 , 0.084 ) | 1.42 ( 1.25 , 1.62 ) | 1 | 11.6 ( 8.3 , 18.8 ) |
| London | 0.072 ( 0.046 , 0.099 ) | 1.52 ( 1.32 , 1.75 ) | 1 | 9.7 ( 7.0 , 14.9 ) |
| North West | 0.070 ( 0.054 , 0.088 ) | 1.51 ( 1.38 , 1.65 ) | 1 | 9.8 ( 7.9 , 12.9 ) |
| North East | 0.058 ( 0.030 , 0.089 ) | 1.41 ( 1.20 , 1.66 ) | 1 | 12.0 ( 7.8 , 22.8 ) |
| South East | 0.060 ( 0.039 , 0.082 ) | 1.42 ( 1.26 , 1.60 ) | 1 | 11.6 ( 8.4 , 18.0 ) |
| South West | 0.065 ( 0.032 , 0.103 ) | 1.46 ( 1.22 , 1.78 ) | 1 | 10.7 ( 6.8 , 21.4 ) |
| Yorkshire | 0.046 ( 0.023 , 0.070 ) | 1.32 ( 1.16 , 1.50 ) | 1 | 15.1 ( 9.9 , 29.5 ) |

**Table S2.**
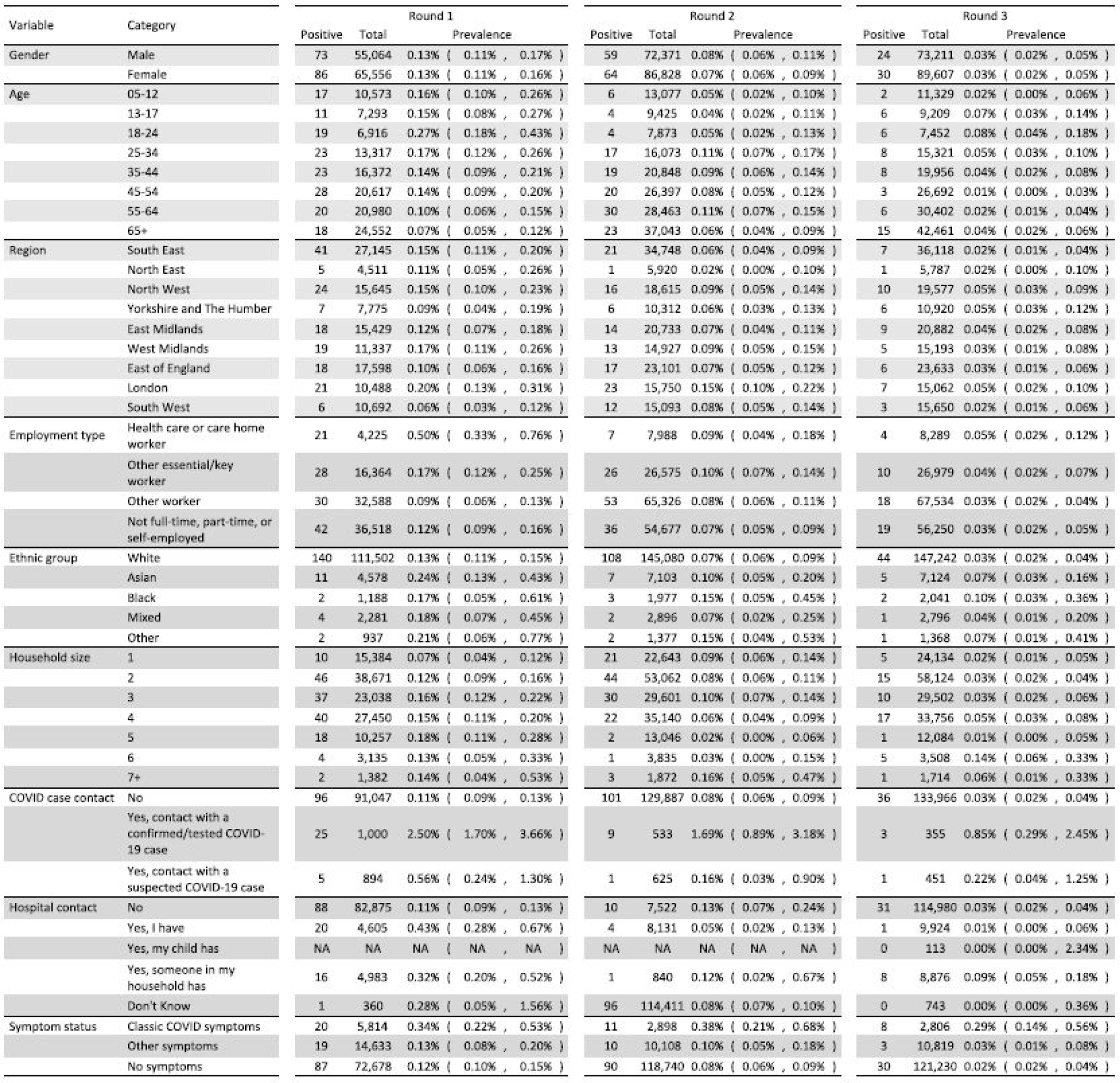
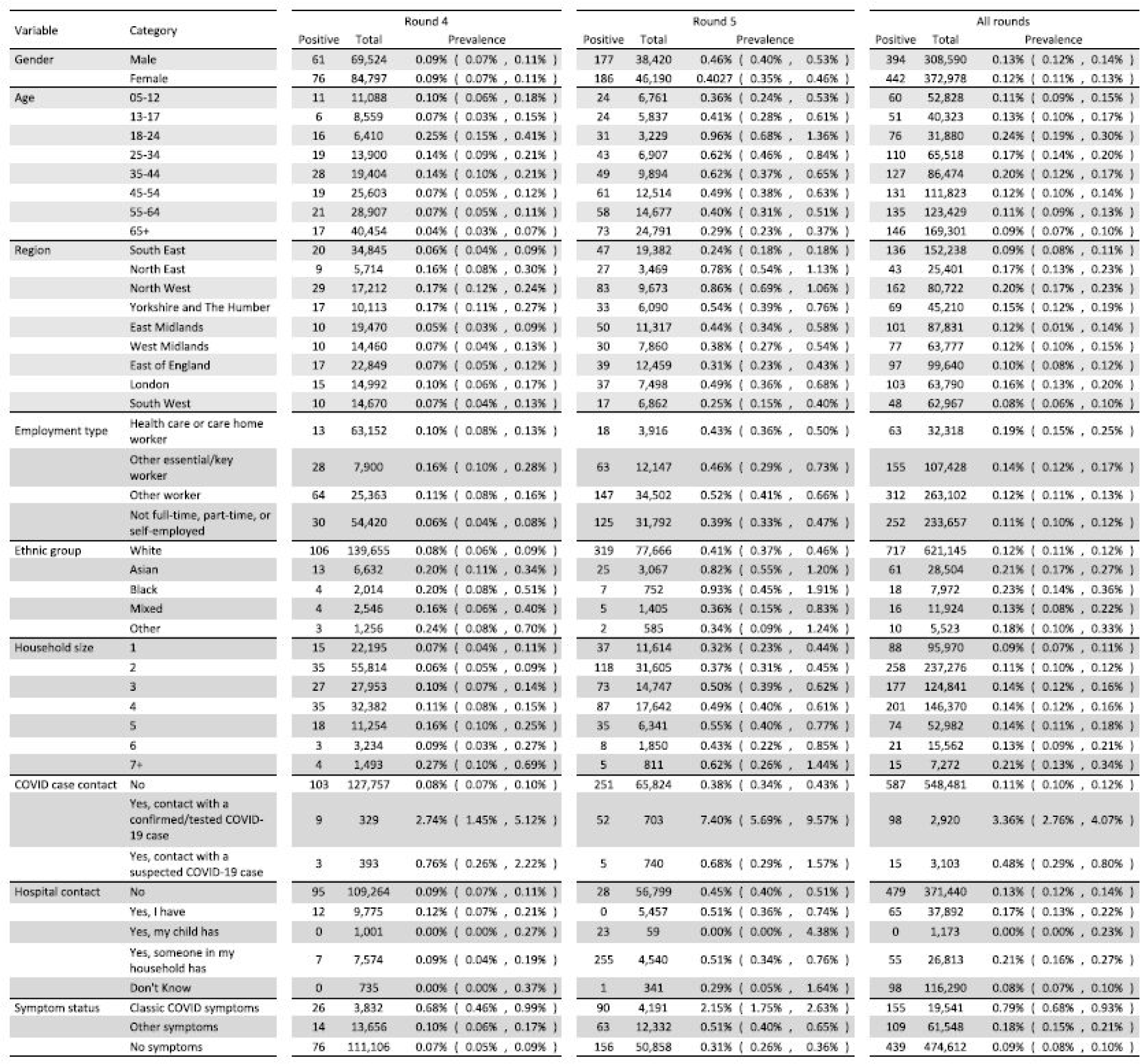
Raw prevalence of swab-positivity by variable and category for REACT-1.

**Figure S1.**
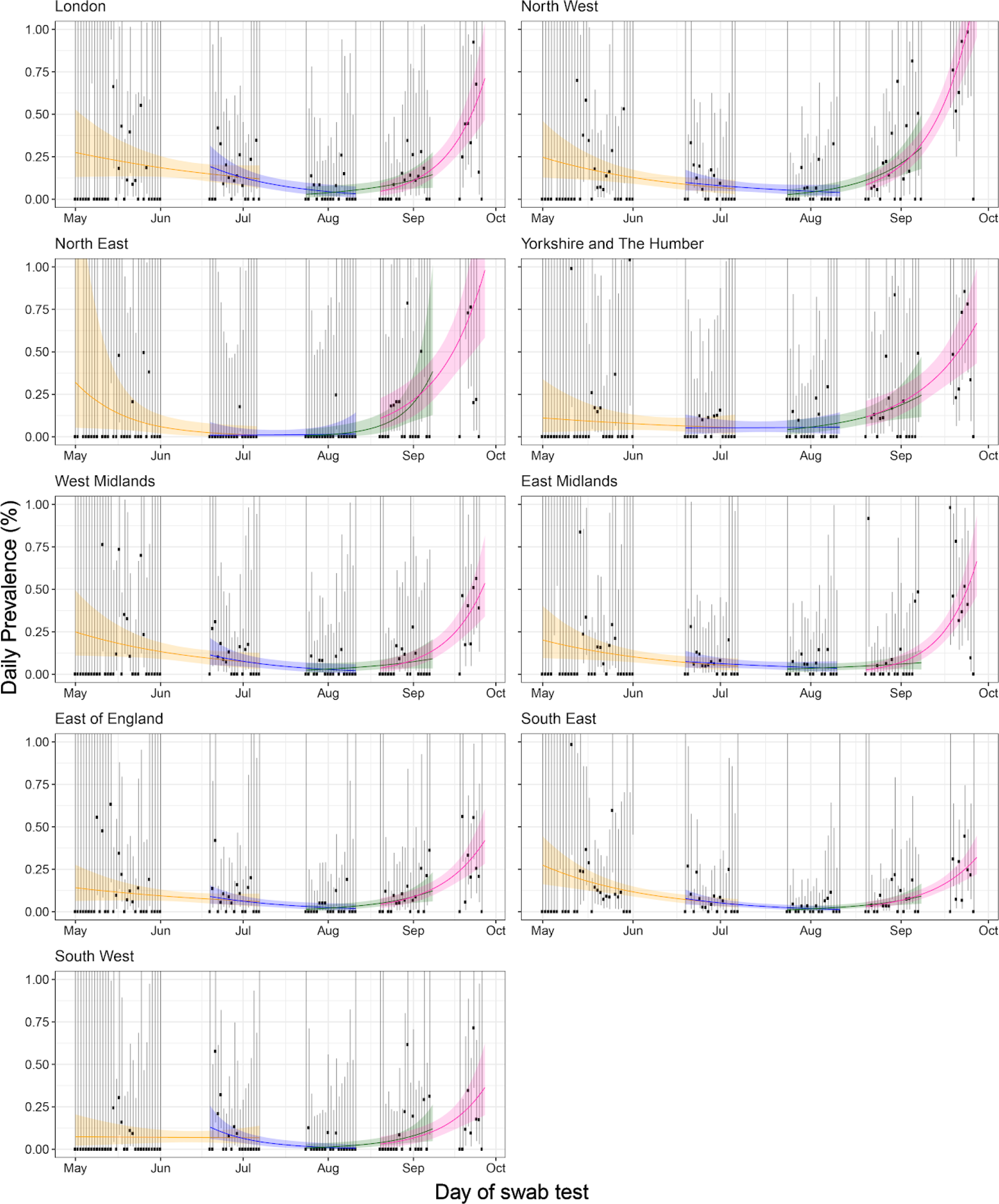
Constant growth rate models fit to REACT-1 data regions for sequential rounds. Models fit to REACT-1 data for sequential rounds 1 and 2 (yellow), 2 and 3 (blue), 3 and 4 (green) and 4 and 5 (pink). Vertical lines show 95% confidence intervals for observed prevalence (black points). Shaded regions show 95% posterior credibility intervals for growth models.

**Figure S2.**
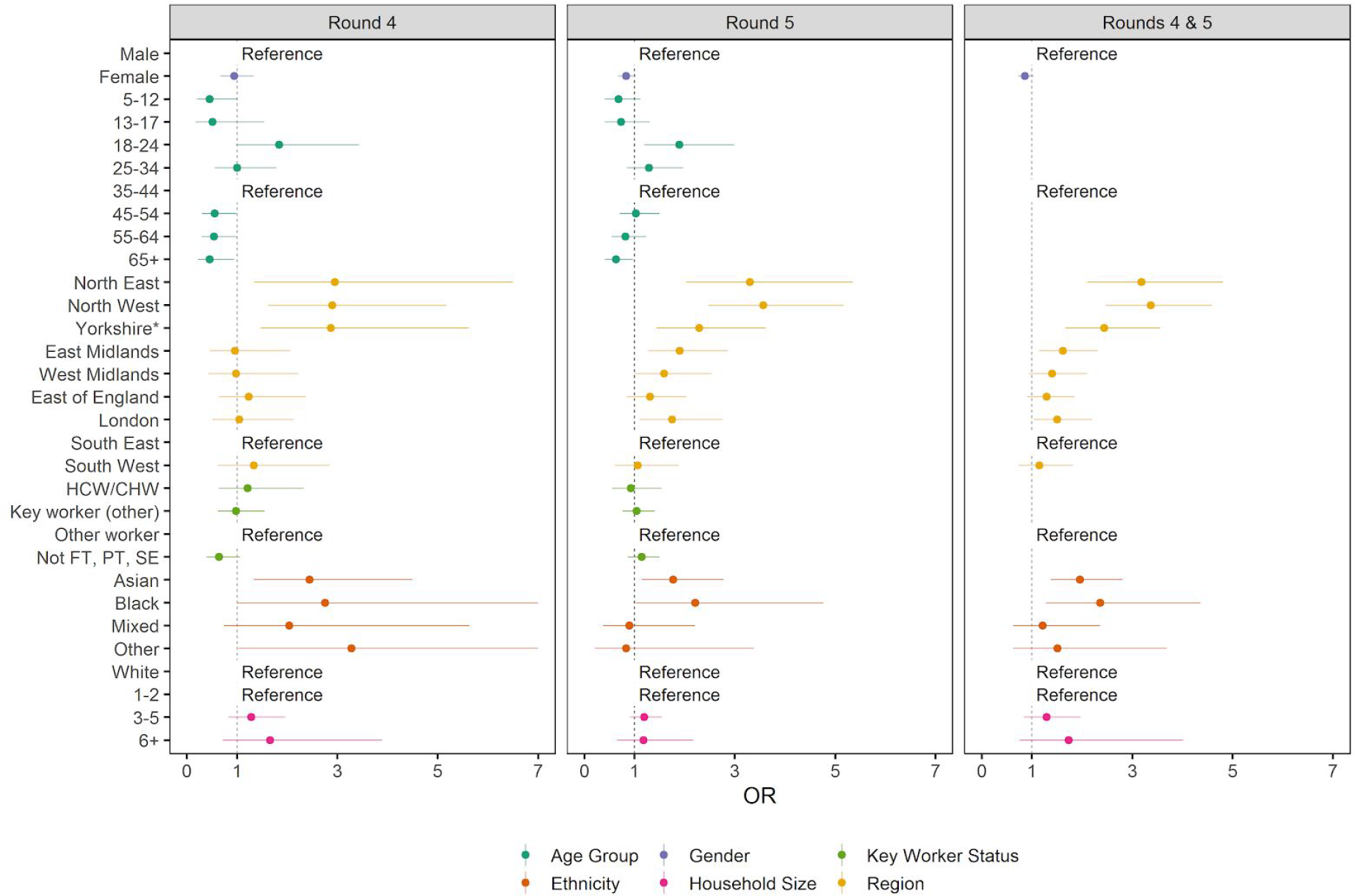
Swab-positivity odds ratios for key epidemiological characteristics. Odds ratios and 95% confidence intervals for epidemiological characteristics (gender, age group, region, key worker status, ethnicity, and household size). Odds ratios were obtained by performing multivariable logistic regression analysis of REACT 1 data from round 4, round 5, and rounds 4 and 5 together. The model for rounds 4 and 5 together was jointly adjusted for round, gender, age, region, key worker status, ethnicity, and household size with interaction terms for age by round and key worker status by round. The rightmost plot (Rounds 4 & 5) only shows odds ratios for variables that were fitted without an interaction term. Within the plot, “Reference” indicates the category that was used as the reference group. * Yorkshire and The Humber. HCW / CHW = health care worker / care home worker, Not PT, FT, SE = Not part-time, full-time, or self-employed.

## References

1. Dong E, Du H, Gardner L. An interactive web-based dashboard to track COVID-19 in real time. Lancet Infect Dis. 2020;20: 533–534.

2. Riley S, Atchison C, Ashby D, Donnelly CA, Barclay W, Cooke G, et al. REal-time Assessment of Community Transmission (REACT) of SARS-CoV-2 virus: Study protocol. Wellcome Open Research. 2020. p. 200. doi: 10.12688/wellcomeopenres.16228.1

3. Riley S, Ainslie KEC, Eales O, Jeffrey B, Walters CE, Atchison CJ, et al. Community prevalence of SARS-CoV-2 virus in England during May 2020: REACT study. Infectious Diseases (except HIV/AIDS). medRxiv; 2020. doi: 10.1101/2020.07.10.20150524

4. Riley S, Ainslie KEC, Eales O, Walters CE, Wang H, Atchison CJ, et al. Resurgence of SARS-CoV-2 in England: detection by community antigen surveillance. medRxiv. 2020; 2020.09.11.20192492.

5. Hoffman MD, Gelman A. The No-U-Turn Sampler: Adaptively Setting Path Lengths in Hamiltonian Monte Carlo. arXiv [stat.CO]. 2011. Available: http://arxiv.org/abs/1111.4246

6. Bi Q, Wu Y, Mei S, Ye C, Zou X, Zhang Z, et al. Epidemiology and Transmission of COVID-19 in Shenzhen China: Analysis of 391 cases and 1,286 of their close contacts. Infectious Diseases (except HIV/AIDS). medRxiv; 2020. doi: 10.1101/2020.03.03.20028423

7. Böger B, Fachi MM, Vilhena RO, Cobre AF, Tonin FS, Pontarolo R. Systematic review with meta-analysis of the accuracy of diagnostic tests for COVID-19. Am J Infect Control. 2020. doi: 10.1016/j.ajic.2020.07.011

8. Verity R, Okell LC, Dorigatti I, Winskill P, Whittaker C, Imai N, et al. Estimates of the severity of coronavirus disease 2019: a model-based analysis. Lancet Infect Dis. 2020;20: 669–677.

9. Christensen H, Turner K, Trickey A, Booton RD, Hemani G, Nixon E, et al. COVID-19 transmission in a university setting: a rapid review of modelling studies. Health Policy. medRxiv; 2020. doi: 10.1101/2020.09.07.20189688

10. Henretty N, Cooper J. Coronavirus (COVID-19) Infection Survey pilot - Office for National Statistics. Office for National Statistics; 2020 Jul. Available: https://www.ons.gov.uk/peoplepopulationandcommunity/healthandsocialcare/conditionsanddiseases/bulletins/coronaviruscovid19infectionsurveypilot/31july2020

11. UK Government. UK government Covid-19 dashboard. In: UK government Covid-19 dashboard [Internet]. Available: https://coronavirus.data.gov.uk/

12. Cabinet Office. Coronavirus (COVID-19): What has changed –22 September. In: GOV.UK [Internet]. 22 Sep 2020 [cited 30 Sep 2020]. Available: https://www.gov.uk/government/news/coronavirus-covid-19-what-has-changed-22-september

13. Riley S, Ainslie KEC, Eales O, Walters CE, Wang H, Atchison CJ, et al. Transient dynamics of SARS-CoV-2 as England exited national lockdown. medRxiv. 2020. Available: https://www.medrxiv.org/content/10.1101/2020.08.05.20169078v1?rss=1%22

